# Barriers and facilitators along the Hepatitis C reinfection journey: Qualitative interview study

**DOI:** 10.1101/2025.11.07.25339747

**Authors:** Avelie Stuart, Cristina Santamaria-Plaza, Matthew Hibbert, Ruth Simmons, Leila Reid, Mark Roberts, Jackie Moore, Monica Desai

**Affiliations:** Blood Safety, Hepatitis, STIs and HIV Division, UK Health Security Agency, Colindale London UK NW9 5EQ; Hepatitis C Trust, 72 Weston St, London SE1 3QH

**Keywords:** hepatitis C virus reinfection, hepatitis C elimination, prevention, harm reduction, COM-B, people who inject drugs, direct acting antivirals

## Abstract

Among people who have experienced hepatitis C virus (HCV) reinfection, we examined the barriers and facilitators to preventing (re)infection with HCV, to seeking testing and treatment, and for prospective prevention of further reinfection. Remote semi-structured interviews (January-August 2024) were conducted with participants recruited via the Hepatitis C Trust and the National Health Service in England. The data were thematically analysed using the COM-B model, exploring how capability, opportunity, and motivation affect behaviour during participants’ journeys with (re)infection-testing-treatment. Six participants were interviewed including 1 female, 1 person who acquired HCV via sex, 1 person who received care in prison, and 5 people who injected drugs. Some people who inject drugs reported not feeling capable of preventing reinfections whilst injecting drugs. Additional capability barriers were lack of knowledge of transmission risks beyond needles/syringes, and knowledge of treatment. During subsequent treatment, participants reported better knowledge of transmission risks and treatment, and that peer support helped motivate them to complete treatment. Participants also reported less stigma from healthcare professionals since their initial infection or treatment. Their future reinfection prevention strategies included opportunity and motivational changes including changing their social circles and environment, prioritising their health, and engaging in volunteering. Therefore, health services are an opportunity for education and behaviour change interventions, including peer support programmes, to support prevention of reinfection. However gaps remain in continuity of care and harm reduction access, which are also needed to enable opportunities to prevent first time infections.

## Introduction

The WHO Global Health Sector Strategy (GHSS) calls for the elimination of blood borne viruses and sexually transmitted infections, including hepatitis C virus (HCV), by 2030 (World Health Organization, 2022). The introduction of Direct Acting Antivirals (DAAs) in 2015, which can cure 95% of hepatitis C infections, and scale-up of testing and DAA treatment is associated with a decline in the prevalence of hepatitis C. For example, in England estimates suggest a 56% reduction of people who are living with chronic hepatitis C since 2015 (Mitchell, 2025). However, reinfection may pose a threat to the gains made in progressing towards the goals for elimination of HCV (Falade-Nwulia et al., 2018) and in maintaining low rates of incidence.

The majority of HCV transmission worldwide is linked to injecting drug use (Nelson et al., 2011). People who have been previously infected with the virus but continue to inject or engage in other risk behaviours are at risk of experiencing hepatitis C reinfection. It is important to understand the drivers of reinfection and barriers and facilitators to engaging in care to inform the development of supportive initiatives to reduce this risk (Mitchell, 2025).

### Population reinfection rates and risk factors

Previous analysis conducted by the UK Health Security Agency (UKHSA) using surveillance datasets found that HCV reinfection among those initially treated in England between 2015-2019 was 7.91 per 100 Person Years (PYs) (Hibbert et al., 2023), although a lower rate has been observed when including more recent years (6.4 per 100 PYs) (Mitchell, 2025). The reinfection rate is highest among current People Who Inject Drugs (22.55 per 100 PYs) and people who were in prison at first treatment (20.42 per 100 PYs) (Hibbert et al., 2023). Women had a significantly reduced risk of reinfection, whereas being of younger age, currently injecting drugs, and receipt of first treatment in prison were significantly associated with increased risk of reinfection. Further international research has found that reinfection is a risk with people who inject drugs and people living with homelessness (Askar et al., 2022; Černoša et al., 2025; Chacón et al., 2024; Fierer et al., 2024). For example, a program for people who inject drugs in Slovenia found that 21.4% of people whose infection was no longer detectable after treatment, later presented with reinfection.

Until DAAs, uptake of HCV testing was low in general and more common in certain contexts such as specialist drug treatment centres (Harris et al., 2014). However, this has changed with the expansion of testing and care programmes to eliminate HCV. Expansion of testing has been linked to prevention of onward transmission of HCV (Bennett et al., 2015; Garvey et al., 2021). However in some contexts, reinfection rates and treatments have risen since the scale up of DAAs, along with a risk that people acquiring a reinfection may not be picked up promptly if they are not frequently re-tested (Černoša et al., 2025; Yeung et al., 2022). Askar et al. (2022) highlighted a concern that the focus on upscaling treatment has taken away attention from adequate harm minimisation efforts such as needle and syringe provision. They also suggested that comprehensive testing in certain settings means that reinfections are being identified at higher rates in some settings more than others, suggesting a need to examine contextual factors.

### Barriers and facilitators to testing, treatment and prevention of reinfection

Qualitative research can complement public health epidemiology research, by examining people’s experiences of barriers and facilitators when seeking testing, treatment or in preventing (re)infection. A common barrier to initial engagement in hepatitis C care is due to the stigma associating hepatitis C with contagion, injecting drug use, or sexual behaviour (Harris et al., 2021; Tam, 2019). Stigma can be enacted by healthcare providers, internalised by people with hepatitis C, anticipated – where people expect unfair treatment, and secondary – entailing judgement or ostracism from family and friends (Tam, 2019). Enacted stigma in the form of prejudice or discrimination from a professional healthcare provider can result in some people receiving poorer healthcare services than others and can reduce people’s willingness to engage with health services. Internalised stigma – associated with feelings of lack of self-esteem and shame – can result in people avoiding seeking help. All stigma can increase stress and worsen mental health (Tam, 2019).

Additional barriers to people’s engagement in hepatitis C care include fear or distrust of hospital settings and procedures, concern about treatment side effects, lack of information, concern about mental health impacts or balancing other aspects of health, amongst other barriers (Amoako et al., 2021; Harris et al., 2021; Harris et al., 2014; Sublette et al., 2015). Facilitators to engagement in hepatitis C care include increasing people’s knowledge and providing integrated interventions through which people can access HCV care alongside additional services and support (Amoako et al., 2021). For instance, people receiving opioid substitution therapy and/or engaged in prison services report increased opportunities and desire to engage in ongoing testing and harm reduction (The Hepatitis C Trust, 2022). Peer support programmes have also been reported to help people engage in care (Jugnarain et al., 2022; Stagg et al., 2019).

There is also concern that new ‘cure is easy’ messaging may complicate reinfection amongst people still at risk, as perceived risks are lowered, people may be less motivated to protect themselves (Yuan et al., 2022). In people co-infected with HIV and acquiring HCV through sexual behaviour, it was found that participants lacked knowledge of how to reduce reinfection risks but also were sometimes unconcerned about reinfection (Marshall et al., 2022). Another consideration is that adequate harm reduction access needs to be made available to people during and after treatment (The Hepatitis C Trust, 2022), otherwise there are insufficient opportunities for them to make harm minimisation choices.

Most of the research cited has been on single infection/treatment experiences or was conducted prior to the upscaling of DAA treatment. Specifically relating to reinfection, a qualitative study with people who were at risk of acquiring hepatitis C again (The Hepatitis C Trust, 2022) found that participants who had been recently treated and were receiving prevention information were personally quite concerned about reinfection. Another interview study with people who inject drugs in Scotland (McEwan et al., 2022) reported that reinfection could be experienced as a ‘double stigma’ due to feeling personal shame and being stigmatised by others for not having prevented reinfection.

### Current study

There are still gaps in knowledge about people who acquire hepatitis C more than once through different routes and in different settings. In addition, many of the barriers and facilitators identified in previous research are about initial hepatitis C infections, there is less qualitative research on reinfection. This interview study was designed to explore barriers and facilitators to testing and treatment occurring across people’s infection-treatment-reinfection journey. We adopt the COM-B model (Michie et al., 2011) to help us analyse people’s capability, opportunity, and motivations to engage in behaviours that prevent (re)infection or to seek testing and treatment in health services. Questions derived from literature about harm reduction, stigma, and testing and treatment experiences are analysed within the COM-B model.

## Method

### Design

Semi-structured interviews were chosen as they allow for an in-depth exploration of individual experiences. Interviews were conducted via telephone calls (n=5) or via Microsoft Teams (n=1), to enable arrangement of the interview at a time and place that suits the participant. This remote method can enhance the anonymity and comfort of patients with hepatitis C reinfection (McEwan, 2023, personal communication). The study setting is England.

### Participants

Of six participants, most were male (one female), three were white British, one was Black African and two did not report their ethnicity. Two participants ages were between 30-39, three were 40-49, and one participant was in the range of 50-59. Three participants were living in north-east England, two in the West Midlands, and one lived in London.

### Materials

The interview topic guide was developed using the COM-B model, widely used in health research to understand barriers and facilitators to target behaviours (Michie et al., 2011). The COM-B contains 3 psycho-social domains: Capability (physical and psychological), Motivation (automatic and reflective), and Opportunity (social and physical). After asking questions about demographics and how they initially acquired hepatitis C, questions to reflect COM-B were asked in relation to behaviours: initial and subsequent prevention of hepatitis C infection, seeking testing and treatment, and prospective prevention of reinfection (see Table 1 for example questions). To follow up previous literature, we included questions about knowledge about the risks of reinfection (an aspect of psychological capability), availability of harm-reduction options (e.g. needle and syringe provision, an aspect of physical opportunity). Questions about stigma were included as an aspect of social opportunity, as being stigmatised by others impacts on people’s opportunities to engage in prevention, testing and treatment.

**Table 1.** Example topic guide questions by COM-B component. See supplementary materials for full topic guide.

| COM-B Component | Example Questions |
| --- | --- |
| Capability:<br>Physical or<br>Psychological | (First infection) What did you know about hepatitis C at the time?<br>(After treatment) Did you know it was possible to get hepatitis C again?<br>Did you feel able to prevent yourself from getting hepatitis C again? |
| Opportunity:<br>Physical or<br>Social | (First infection and after reinfection) Who did you talk to about having hepatitis C?<br>(After initial treatment) Where and how often were you being tested?<br>Have you experienced any stigma or discrimination against you because of your hepatitis C status, and (if relevant) injecting drug use? |
| Motivation:<br>Automatic or<br>Reflective<br>mechanisms | (First infection and after reinfection) How did you feel about having hepatitis C?<br>(After treatment) Did you want to keep doing tests?<br>(After reinfection) Did you want further treatment? |

### Ethical statement

NHS Research Ethics Committee and Health Research Authority approval was gained for the recruitment of NHS patients (IRAS: 331334). [anonymised for review] research ethics and governance approval was obtained for recruitment via the Hepatitis C Trust (ID: 553). All participants gave informed consent, and participation was anonymised after completion of transcription.

### Public and patient involvement

Staff and volunteers from the HCT were consulted on the study procedure and topic guide. The topic guide was piloted with one person with lived experience of hepatitis C reinfection. From the feedback we changed informed consent procedures to verbal, simplified language, added questions about exposure to HCV and the type of harm reduction behaviour to enquire about, and created a shorter topic guide for time-constrained participants.

### Eligibility criteria

All participants were adults (18+) and had completed treatment at least once before receiving new subsequent positive PCR RNA tests and further course(s) of treatment (similar to criteria in Hibbert et al., 2023). Due to this requirement to determine whether the case is a reinfection, participants all received their initial treatment in England.

### Procedure

Participants were recruited by the Hepatitis C Trust, a national charity that provides peer-led interventions, or the National Health Service (NHS) in England. The NHS sites were selected based on prevalence of hepatitis C reinfection and varying composition of patients in their populations (e.g., individuals’ who have been in prison, gay, bisexual or other men who have sex with men [GBMSM]) (Hibbert et al., 2023). NHS and HCT staff phoned people known to have acquired hepatitis C more than once to ask permission to share contact details with the research team. People who agreed then had minimal information shared with the research team, who conducted the interviews.

Information and consent were provided verbally to protect against accidental disclosure of hepatitis C status. Consent was recorded separately from the interview recordings. Interviews were audio recorded for transcription purposes. Participants were compensated with a £20 Love2Shop e-voucher.

Interviews started January 2024 and were planned to continue until saturation was reached (15-20 participants), but healthcare professionals and the interviewers were finding it hard to contact potential participants. Thus, recruitment ended in August 2024.

### Reflexivity

The first and third authors were the interviewers, both have PhDs in Psychology and previous experience conducting qualitative interviews, but do not have lived experience of hepatitis C. Initial contact was made by HCT peers or NHS healthcare professionals who were known to the participants. The second author analysed the data and had previous experience conducting qualitative research but was new to hepatitis C.

### Data analysis approach

The smaller sample size presented an opportunity to maximise an in-depth exploration of individual patterns of barriers and facilitators. Rather than thematic saturation, we considered information power (Malterud et al., 2016). Given the specificity of the sample (people who had been treated for multiple hepatitis C infections), and a consideration of individuals experiences at different time points in their infection and treatment journey, we garnered greater information power with a smaller sample (Malterud et al., 2016).

The method of analysis was adapted from combining thematic analysis (Braun & Clarke, 2006) with a life history perspective (Harris & Rhodes, 2018), involving: 1) familiarisation and development of coding plan, 2) inductively coding each participant’s accounts of barriers and facilitators at each time point: initial infections and treatment, reinfection(s) and subsequent treatment, and prospective prevention of future reinfection, 3) deductively assigned codes to the components of the COM-B model, 4) grouping commonly reported experiences across participants to generate themes, 5) discussion within the team led to revision of codes and themes, 6) finalising themes and writing the report with example quotes. Separately, we also summarise participants discussions of the wider impacts of social stigma.

## Results

### Overview of barriers and facilitators

Table 2 lists each participant’s reported barriers and facilitators at each time point alongside the relevant COM-B component for the target behaviour. The time points are separated into: 1) initial infection and first treatment, 2) past reinfections and subsequent treatments, and 3) future prevention of reinfection.

**Table 2.**
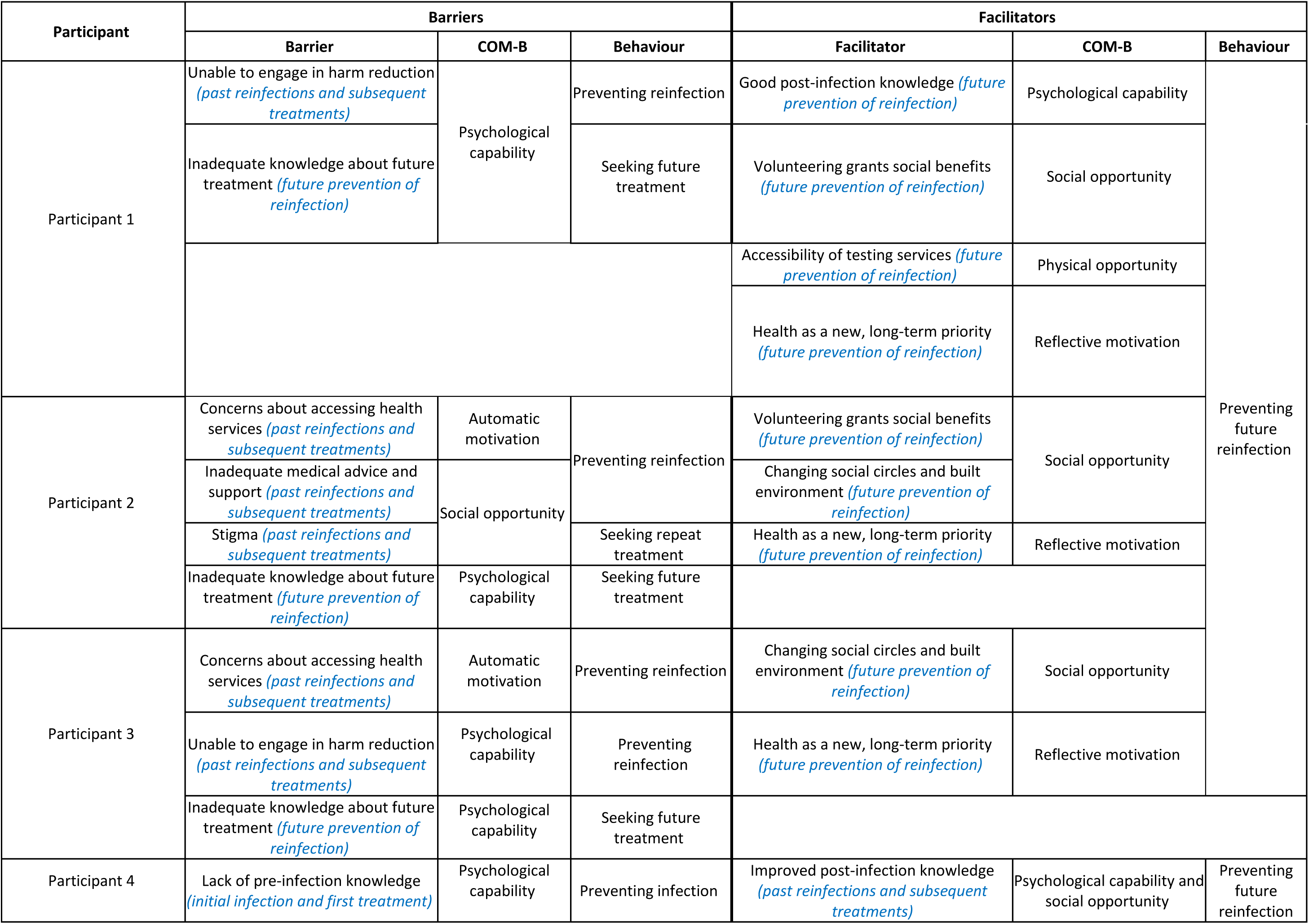
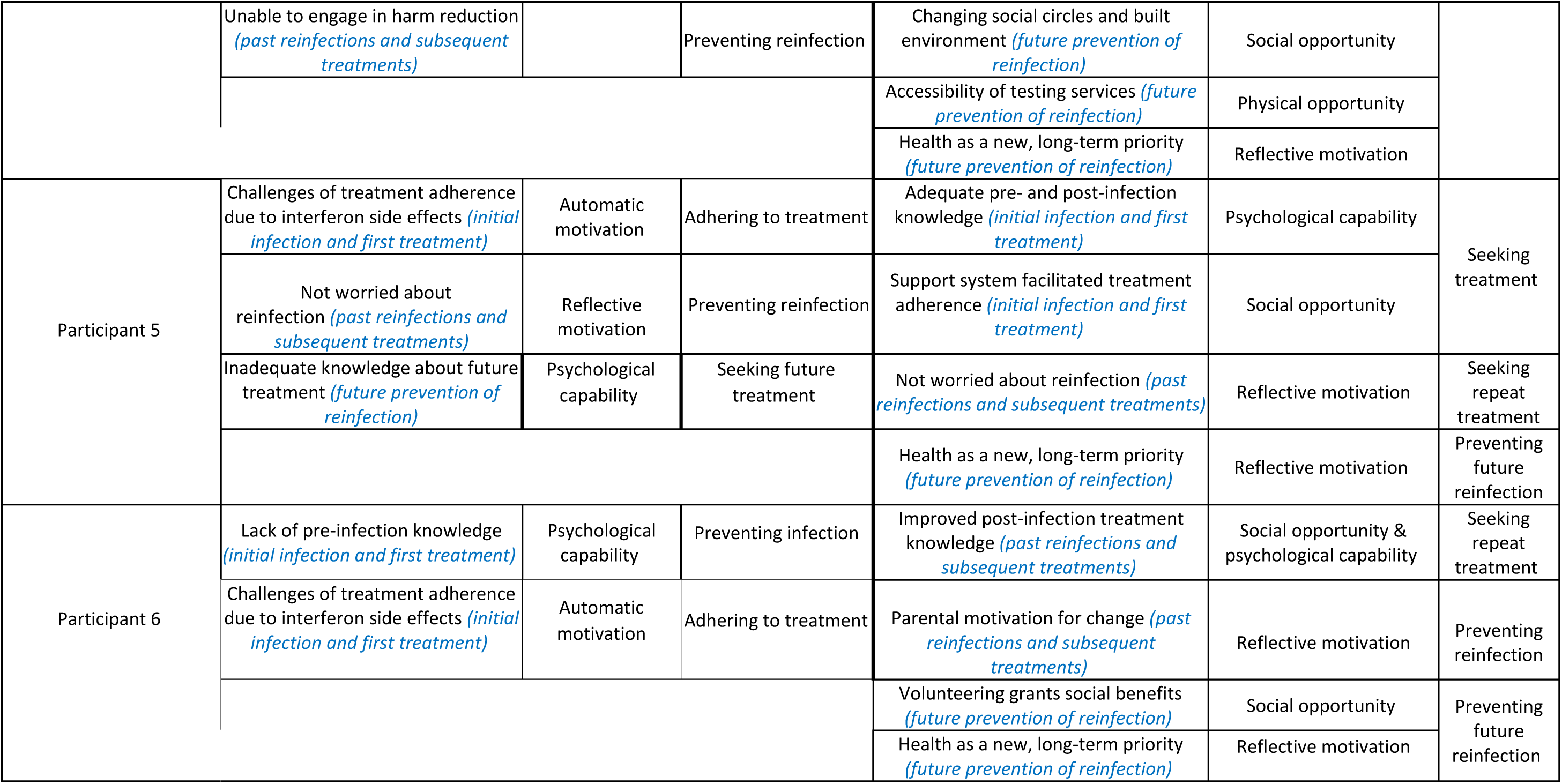
List of barriers and facilitators experienced at different timepoints by each participant.

The results herein are described in order of timepoint, barriers/facilitators, behaviour, and then the theme.

### Initial infections and first treatment

Five participants initially acquired hepatitis C through non-sterile injecting equipment. One participant, who acquired hepatitis C through sexual contact between men, was diagnosed through regular testing during their HIV care. Three participants were tested by an HCT peer worker, who visited participants in their homes or community. One participant was diagnosed in drug treatment services, and another was tested and treated in prison. Two participants had interferon as their first method of treatment (the treatment regime that preceded DAAs and was less effective/tolerable), followed by DAAs for reinfection(s); all other participants only received DAA treatment.

Barriers and facilitators were reported when experiencing a positive hepatitis C test and corresponding treatment for the first time.

Behaviour: Preventing hepatitis C infection

*Participant 4 & 6: Lack of pre-infection knowledge (psychological capability, barrier)*

Prior to acquiring hepatitis C for the first time, both participants expressed that they had limited knowledge on hepatitis C and how it was transmitted. Participant four was only aware that HCV was transmitted through needles. Participant six initially thought HCV was the same thing as HIV and was scared when they received the first diagnosis. In the past, they said there was information available, but it was not as widely promoted or accessible as it is today:

> *[…] when I first found out early because I had no knowledge of it, I was expecting sort of like ohh God, I’ve got [AIDS] or something like that […] And I was in an emotional mess thinking it’s gonna kill us off this sort of thing, cause I had no knowledge about it, it wasn’t, there was information out if you wanted it, if you weren’t bothered when you were going to drug clinics, they weren’t like pushing for tests and things the way they are now, there was no sort of incentives […]*

Participant six also said that it would have been more impactful and beneficial to have prevention information delivered through more engaging means (e.g. hearing from a peer).

Behaviour: Adhering to treatment

*Participant 5 and 6: Challenges of treatment adherence due to interferon side effects (automatic motivation, barrier)*

Participants five and six found that prior treatment with interferon negatively impacted their physical and mental well-being, and for participant five it affected their willingness to receive future treatment. However, both were able to continue through the treatment process. Participant five found the experience cumbersome due to the number of injections required, whereas participant six said the side effects significantly impacted their ability to function in their day-to-day life:

> *[…] [The interferon] crippled me. It absolutely wiped me out […] I was like a broken bloody wheel. I couldn’t function or anything. I carried on with the treatment as rough as it was, and I think me weight went down from being like 12/13 stone to 10…10 stone if I was lucky*.

Conversely, the rest of the participants who had DAAs did not report similar challenges.

Behaviour: Seeking treatment

*Participant 5: Adequate pre- and post-infection knowledge (psychological capability, facilitator)*

Prior to their first infection, participant five had some knowledge of hepatitis C and was aware that it affected the liver. This knowledge prompted them to seek treatment; the participant said that they did not need to know much about how hepatitis C was transmitted to take action:

> *Not that much. I knew that it affected your liver. Um, and that treat [untreated?]*.
>
> *You have problems, with your liver. And that was enough for me. That was enough information for me to actually do something about it*.

They also confirmed that they did not need additional information in the future.

*Participant 5: Support system facilitated treatment adherence (social opportunity, facilitator)*

Throughout their first (and subsequent) treatment, participant five said they received support from their family and other members in their social circle, even when they were receiving interferon, which was more visible:

> *I have had support and all around. And I also had supporting family members as well. Well, thank you. Very good […] they’ve always been helpful. Well useful as well […] what I think is when I was given injection every week, you have to put the medication in the fridge. You have to [unclear] for the medicine in the fridge […] once it was a tablet nobody needed to know. But at that time, you needed to. Yeah. What’s this? What is this thing in the fridge? […] They were supportive*.

They also expressed having experienced no stigma from NHS staff who helped create a positive and supportive environment. Having this social support was described as important for maintaining treatment.

### Past reinfections and subsequent treatments

After initial infections and treatments, the participants had reacquired hepatitis C and received treatment for reinfection. We refer to these as past reinfections and repeat treatments in the following barriers and facilitators.

Behaviour: Preventing reinfection

*Participant 1, 3 & 4: Unable to engage in harm reduction (psychological capability, barrier)*

A barrier to preventing reinfection was not being able to engage in harm reduction whilst using substances. Participants report this as a personal capability issue (hence we have coded it as such), but lack of access to harm reduction should be considered in the interpretation of this finding. For example, participant one said that engaging in substance use impacted their ability to prioritise their personal health and the use of sterile injecting equipment. The participant described “relapsing” for a short period of time where they said they were not being cautious or, taking precautions due to mental health:

> *[…] it [reinfection] wasn’t on my mind. And so, I wasn’t thinking straight at the time, do you know what I mean? I didn’t take any precautions at the time […] And mental health was really bad and stuff*

Participant three resumed injecting drugs after a few years, and despite showing awareness on how to minimise harm, they were infected with hepatitis C again:

> *I thought I didn’t realise the most, the most likely way to catch it is through everyone sucking water out the same… I’ve caught it again. Uh, it’s probably been the same way. But I honestly don’t know how I caught it this time. Cause I was being dead dead careful*.

After their first infection, participant four stopped injecting drugs for 15 years. However, they reported starting to use drugs again. Despite trying to be cautious, they were reinfected with hepatitis C. They also expressed uncertainty about when they were reinfected during that time due to inconsistent test results from different testing methods, however the return to injecting drug use is the reason they were tested:

> *I thought I’d kept myself safe […] I started taking cannabis now and then and I got to the stage where I didn’t really know what I was doing […] I thought I was all clean and safe and everything. The second test was more just a, just an in case of thing, but I ended up finding out I had it again […] I think I might have used someone else’s pin or dirty water again, I’ve been pretty oblivious to what I was doing […] For [many years]. We hadn’t injected for, and then we started doing it all the time and that’s the reason we even got the check*.

*Participant 2 & 3: Concerns about accessing health services (automatic motivation, barrier)*

Participant two and three faced anxiety related barriers when accessing health services. After their second round of treatment and multiple incarcerations, participant two was engaging in injecting drug use and wanted to enter drug treatment rehabilitation. However, personal fears and beliefs about detoxification, and not viewing their health as a priority, prevented them from attending. For the participant, this meant that they felt they were still putting themselves at risk of acquiring hepatitis C.:

> *[…] And I really wanted it, but I was scared. And she [support worker] got me a room. She got me a bed and I didn’t go. But then I just she, she kept trying and trying and trying man. And one day I’d give in and I’ll was just like, “Yeah, I’m ready. Get me.” She was that worried about me, she used to come to the squat just to bring my food and that and check to see if I was still alive […] I was scared of going to rehab […]*

Participant three discussed their anxiety when having to attend hospitals for hepatitis C care, and how certain medication aids in easing that anxiety, but both the participant and the GP were reluctant to prescribe anxiety medication:

> *[…] If I have to go to [hospital] I get dead nervous… I’m walking to the [hospital] like, a nervous wreck. I come back, I’ll [get] panic attacks and everything…I thought my anxiety will get better. I’ve always had bad anxiety, but I thought it would get better. But I think the drugs just masked it. Do you know what I mean? And then when I speak to the doctor about it like, the doctor don’t want to give us anything sort of thing…because I’ll end up addicted to them*.

*Participant 4: Improved post-infection knowledge (psychological capability & social opportunity, facilitator)*

Participant four noted that their hepatitis C transmission knowledge was improved during their first round of treatment. Both the HCT peer support worker and the hepatology staff provided in-depth information on transmission and treatment:

> *[Peer] came around like, and he told us everything about sharing pots and even stupid things like cannabis and crack pipes and things that you got cooked on, you know, right. He told us all the ins and outs to stuff I didn’t really know. You know, I, I was well informed after that to be honest, aye he did a good job, you know […]*

*Participant 6: Parental motivation for change (reflective motivation, facilitator)*

Participant six’s motivation to prevent their past reinfection was driven by their responsibility of caring for their children. For years after clearing hepatitis C the first time, the participant distanced themselves from substance use and removed themselves from situations and friendships that could negatively impact their focus:

> *[…] my children at the time were like, [young]… It was very, very busy life should I say and if it hadn’t been for the [children]. I don’t think I’d be here today to be fair, because that was my that was my focus on keeping myself right […]*

Behaviour: Seeking repeat treatment

*Participant 2: Stigma (social opportunity, barrier)*

Participant two described their hesitancy to get tested while in prison due to stigma and being negatively labelled by others once associated with hepatitis C. This prevented them from engaging with care:

> *Yeah, cause you don’t really want to know because you’re too scared. Cause if you have got it, you think you gonna die, or, if you don’t think you’re gonna die, you think you’re going to get labelled as being dirty like, like having AIDS or HIV or something […] When you’re with ya mates and you’re seeing ‘em at [charity], you don’t want to be going talking to BBV nurses coz then people’ll start talking and saying: “Ooo I bet he’s got hep C he’s been seen with the nurse?” So like it’s fear*.
>
> *It’s all fear. Everything’s about fear these days […] It really stopped me from engaging*.

*Participant 2: Inadequate medical advice and support (social opportunity, barrier)*

In response to a nurse’s comment that they can only be treated once, participant two felt it necessary to lie about their treatment history during their second round of treatment for fear of being denied repeated treatment:

> *The first time I got told that you don’t get treated more than once. It’s very hard. And so, I kind of lied a little bit when I went in the second time. And then when they when they tested me and it come back positive, they got me the treatment. But I was because of what I got told before, I said I hadn’t had the treatment before […] I don’t know if they knew or not, but they just treated it anyway. […] I think she was just trying to get me to not do it again*.

*Participant 6: Improved post-infection treatment knowledge (social opportunity & psychological capability, facilitator)*

The participant was aware of the possibilities of acquiring a reinfection and the roots of transmission. When they were diagnosed again with hepatitis C, they worried about the prospect of interferon treatment again. However, connecting with the HCT peer helped the participant feel informed and capable of managing hepatitis C:

> *[…] after I got me first treatment, I knew the possibilities and the roots of transmission and what have you […] [HCT Trust peer] who was the man who had done the test and I’d spoke to and he put my mind at ease in an instant*.

Behaviour: Preventing reinfection and seeking repeat treatment

*Participant 5: Not concerned with reinfection (reflective motivation, barrier & facilitator)*

Participant five expressed minimal concern about reinfection; we categorised this as a barrier to preventing reinfection, but also a facilitator to seeking repeat treatment:

> *Look it doesn’t bother me. I haven’t really suffered mentally or physically[…]. I just find [treatment] more inconvenient rather than feeling depressed about it […] I just saw it [as one of those things?] because I thought I just have to get through with it. But that’s my personality anyway*.

### Future prevention of reinfection

The participants emphasised preventing future reinfections and discussed prospective barriers and facilitators.

Behaviour: Seeking future treatment

*Participant 1, 2, 3 & 5: Inadequate knowledge about future treatment (psychological capability, barrier)*

A few participants expressed concerns about obtaining future treatments. Participant one was under the impression that the treatment *“isn’t going to keep reacting.”*. Participant three expressed a similar sentiment:

> *…the more you get [hepatitis C], the harder it is to get rid of*

Participant five expressed uncertainty about whether they would acquire liver damage even with successful repeat treatments. It was unclear where participants had obtained these concerns.

Behaviour: Preventing future reinfection

*Participant 1: Good post-infection prevention knowledge (psychological capability, facilitator)*

After undergoing their second round of treatment, participant one felt better equipped to prevent future hepatitis C infections and felt confident in their knowledge to maintain their behaviour:

> *No [I don’t think there is more information people should be given], not really. No. Um. Right, because I think it’s all explained pretty well*.

*Participant 1, 2 & 6: Volunteering grants social benefits (social opportunity, facilitator)*

Three participants shared how volunteering with the HCT had granted them a greater sense of purpose, which encouraged their commitment to adhering to treatment and preventing further reinfection. Participants one and two felt they served as positive social influences for others undergoing testing and treatment for hepatitis C. Being a volunteer has been instrumental for participant six in the period since they stopped using drugs:

> *I think the best thing about it is the general feeling I get inside that I’ve actually been out and I’ve helped somebody and I’ve helped them and they’re going to progress in their life […] So it’s like you’ve saved a life at the end of the day, which is…no drug I’ve ever took can compare to the feeling I get from going out and helping people mind to me it’s amazing like I can’t praise [HCT peer worker] enough because he’s totally changed me life around and me outlook on things and it’s just giving us something to focus on*.

*Participant 2, 3 & 4: Changing social circles and built environment (social opportunity, facilitator)*

Participants two and three found that changing their social circles reduced the opportunity to engage in substance use and, in turn, enabled prevention of future reinfection. For participant two, this shift occurred when they moved from their hometown and entered a rehabilitation clinic after their second round of hepatitis C treatment. Participant three discussed negative social pressures from those in their social circle that still engaged in substance use, so they ceased being friends with them:

> *[…] it’s unreal, like when we got clean and that it was unreal how much people tried to drag us back down into it […] But it’s getting to the point where I’ve literally had to tell people to f off. […] And fall out with people, because of this and they’re not friends anyway*.

For participant four, this behaviour change was openly talking with others which has improved their understanding of transmission risks, how to prevent infections and removal of stigma attached to hepatitis C. They also said they inform others on the risks of using non-sterile drug equipment.

*Participant 1 & 4: Accessibility of testing services (physical opportunity, facilitator)*

Another facilitator was the increased availability of testing services. For participant one, after their first round of treatment, they became an HCT volunteer which granted a physical cue to support the behaviour of preventing hepatitis C reinfection:

> *[…] You know, I put myself on a Cepheid machine last week. Um, you know, and obviously I’m hepatitis free again at the moment […] I haven’t got any plans on catching it a third time, you know? But, you know, I suppose I’m lucky being up here as well, like I said I get to put myself on the machine and, you know, test myself […]*

Participant four also discussed their future intentions to continue testing, which was facilitated by the increased availability of testing services where they collect methadone, acting as a physical cue to maintain regular testing.

*Participant 1, 2, 3, 4, 5 & 6: Health as a new, long-term priority (reflective motivation, facilitator)*

All participants discussed shifting from engaging in behaviour that put them at risk of reinfection, to prioritising their long-term health. Having gone through two rounds of treatment, participant three felt more motivated to prevent further reinfection. Coupled with their experience with sepsis, the participant wanted to seek treatment with methadone to improve their health and to prevent further reinfections via injecting drug use:

> *I ended up with an infection […] it was just a massive wakeup call […] we didn’t get clean straight away. Like […] that was the start, you know like that was about a year ago […] I’m not getting it [hepatitis C] again. I’m not getting it again like I’m totally, definitely cracked the using. Like I’m done with the [unclear] Yes, I really am […]*

Similarly, after their second infection, participant four felt a greater sense of capability to prevent future reinfection. They threw out injecting equipment, distanced themselves from certain people, and collaborated with police to keep the community safe. Finding out about their hepatitis C reinfection was reported to give them a second chance at a positive change.

Participant five discussed that they were aware that engaging in certain sexual activities could put them at a greater risk of reinfection. Recognition that having hepatitis C in older age could lead to poor health outcomes and liver damage, even with treatment, motivated them to consider reducing their engagement in behaviour that puts them at risk of reinfection:

> *There’s a certain act which one does, which is what, what exposes one to, [a] greater probability of catching it […] I think probably because of my age, I’m getting on now, that’s prevent doing that. So then yeah. […] But obviously [all the more?] because the more I think that prevention is better than the cure […] But you are in a much worse state. Were you to have Hep C, you know, in your older ages and that may have an impact on how one behaves*.

Participant one expressed similar motivations, however, the focus was more so on creating a new life. This change occurred after completing their second round of treatment. Participant six echoed this sentiment by framing their hepatitis C diagnosis to inform a renewed focus on health and well-being, resulting in a more optimistic outlook– motivating them to build new relationships and work towards improving their health:

> *I can actually, it’s stupid and it’s crazy as this might sound having hepatitis C has been the best thing that’s happened to me […] I’m actually thankful for having hepatitis C because of the route it’s took m[y] life now on a different, completely different road to the […] road of chaos and destruction I was already on. It sort of took me away from that. Erm, it’s giving me something else to focus on. […]*

### The broader challenge of stigma

Apart from participant two, the other participants did not report stigma as a barrier to seeking testing or treatment, but they discussed how stigma impacts general wellbeing and their connections to social communities. Participant one experienced multiple instances where they were treated differently because of their hepatitis C status, which impacted their mental health. They were careful to share their hepatitis C status with people out of fear of being labelled as “dirty” or being further discriminated against by friends and family.

As a HCT volunteer, participant two reflected on the past stigma associated with testing for hepatitis C and said that they are reducing stigma by helping to provide accessible services. Participant three experienced stigma from their local community who they felt stereotyped people who inject drugs. Participant four described being selective about disclosing their hepatitis C status, primarily sharing it with close friends to reduce onward transmission. The participant’s fear of judgement impacted their social interactions and support networks. This same sentiment was expressed by participant six who faced varied reactions across social groups:

> *[…] yes, from people who don’t, don’t understand it and don’t have any knowledge about it, but like I say, I was a bit clueless myself back in the day, but if someone hears hepatitis C, I mean if in certain circles, there’s no judgment, but outside of that little chaotic lifestyle you’re living in, the public do look at things and think ‘aw he’s got hepatitis’ and the obviously they’re looking at it like it’s bloody HIV or something because they’ve got no, no knowledge of it*.

Reduction of stigma against people who inject drugs was noted by participant six as a positive wider change that contributes to a more supportive atmosphere in health services that provide drug treatment (not only with hepatitis C care):

> *Nowadays, I find it, it’s all judgment free. Nobody cares anymore. Everybody that everyone sort of knows what you’re there for back when I was 18 year old or something, I felt personally that there was a there’s a lot of judgment of the pharmacies themselves […] it’s not like that anymore I think everybody’s come to the realisation that drugs are an epidemic*.

## Discussion

### Findings on past infections

Barriers for people’s initial and reinfections with hepatitis C occurred in psychological capability (lack of transmission knowledge, unable to engage in harm reduction), automatic motivation (challenges adhering to interferon treatment, concerns about accessing health services) and social opportunity domains (inadequate medical advice/support, stigma). Whilst lack of harm reduction provision was not explicitly stated, it should be considered a key factor in any feelings of being unable to engage in harm reduction. Facilitators were in psychological capability (knowledge of transmission), social opportunity (support systems facilitated knowledge and treatment adherence), and reflective motivation (parental responsibility). Some of these are similar to previous research on first hepatitis C infections (e.g. knowledge barriers, Amoako et al., 2021; Harris et al., 2014; Treloar & Rhodes, 2009), however, we also found new facilitators in these interviews.

Compared to their first infections, participants reported positive experiences of subsequent engagement in care, less stigma and discrimination, access to care in ways that reduced anxieties about conventional hospital appointments, few treatment side effects, and more information about transmission and treatment from healthcare providers and peers.

For three participants, hepatitis C was detected while receiving other care (e.g. HIV treatment or opioid substitution therapy) in a setting where testing is offered regularly. Further, the participants who had their reinfection identified through peer visitors may not have otherwise been tested; these are positive outcomes. However, this study highlighted that gaps can exist in transitions between services (e.g. good availability of health services in prison, but lack of post-release care and prevention – as identified in Carson et al., 2022; Walker et al., 2024) and that harm reduction remains a key issue for preventing (re)infection (Askar et al., 2022).

### Future prevention after reinfection

For future prevention of reinfection, one barrier was identified in psychological capability - some participants expressed doubts about their eligibility for, or the efficacy of, repeated DAA treatment. Fontaine et al. (2023) found a similar belief but related to the cost of DAAs. There is no clear characterisation of treatment resistance (European Association For The Study Of The Liver, 2011; UK Health Security Agency., 2024), and we suggest caution in giving patients information about possible treatment resistance.

These interviews were optimistic in tone, thus multiple prospective prevention facilitators were identified. Participants attributed greater psychological capability (knowledge of transmission risks beyond needles/syringes), social opportunity (volunteering, changing social circles and environment), physical opportunity (accessibility of testing), and reflective motivations (health as new long-term priority). For people who previously injected drugs, these facilitators for infection prevention were tied to ceasing drug use and changing social circles, volunteering, or redefining their personal health goals. This reflects evidence from the social identity model of recovery (Best et al., 2016), whereby overcoming substance addiction involves supplanting a stigmatised “addict identity” with a new positive social identity and community (see also Amoako et al., 2021; Lafferty et al., 2020; Treloar & Rhodes, 2009). Unlike in Lafferty et al. (2020), which involved people currently in prison, participants in this study viewed this identity change as permanent. The role of peers in providing information and social support was emphasised by participants for enabling them to make these changes (also found in The Hepatitis C Trust, 2022).

We also found, similar to previous research (Brothers et al., 2023; Marshall et al., 2022), that the participant who acquired HCV through sex, and was co-infected with HIV, was not very concerned about acquiring a reinfection and felt that the treatment was not onerous to retake as needed; however they did express concern that as they get older their health may be impacted by reinfections.

Stigma was noted as a persistent generalised challenge despite fewer reports of personal encounters than in the past. As in previous research, participants reported secondary stigma (from community, family and friends) and internalised stigma (in feelings of shame and not seeking help) (Tam, 2019; Treloar & Rhodes, 2009). These could impact on people’s health and wellbeing (Harris et al., 2021; Tam, 2019) even if enacted stigma is reduced, especially if people have additional stigmatised identities associated with injecting drug use (Lekas et al., 2011).

### Strengths and limitations

A limitation is that participants were engaged in services and peer support and may feel more positive and well informed than people who have not received support/services. However, these findings are an encouraging indication of the benefits that engagement in care can bring.

Additionally, we were still able to identify remaining gaps in care where care can be improved. Whilst there are limitations on what claims we can make based on the sample size, we did demonstrate in the discussion that there are similarities between our findings and other literature. This sample also allowed opportunity to expand on individual differences, and how unique barriers and facilitators can be encountered. Another limitation was that interviewees could not always recall dates and details of testing and treatment; supplementation with patient records may have helped. An additional strength is the use of the COM-B model to structure the barriers and facilitators, which enables an understanding of the underlying behavioural mechanisms and how interventions might target these – as discussed next.

### Recommendations for intervention and policy

Based on the COM-B domains and barriers and facilitators, in Table 3 we suggest intervention types and social/policy implications (Michie et al., 2011; West et al., 2020). Future interventions aiming to reduce the occurrence of HCV reinfection, should consider the specific interventions in relation to the domain of the COM-B model they would like to address.

**Table 3.** Suggested intervention functions and policy categories from the main barriers and facilitators by domain. This is not a complete list of all options, see (Michie et al., 2011).

| Barriers according to COM-B component | Intervention functions | Policy area |
| --- | --- | --- |
| Psychological capability (e.g. knowledge of treatment) | Education, training, enablement | Communication/marketing, guidelines, service provision |
| Social opportunity (e.g. support systems, stigma) | Environmental restructuring, Enablement | Guidelines, Environmental/social planning, service provision |
| Automatic motivation (e.g. anxiety about health services) | Persuasion, incentives, environmental restructuring, modelling, enablement | Communication/marketing, Guidelines, Service provision, Environmental/social planning |
| Reflective motivation (e.g. health as a priority) | Education, persuasion, incentives | Communication/marketing, Guidelines, Service provision |

Whilst classically, education and training can be useful for improving knowledge, enablement can also support knowledge improvements and reduce automatic motivation barriers such as anxiety. Enablement refers to a more encompassing behavioural intervention that equips people with the means to change behaviour (and can include other components such as education and improvements to social and physical environments). Peer programmes such as the support provided by the HCT fits such a definition of enablement; this study suggests benefits for knowledge and self-efficacy for volunteers and receivers of peer support (see also Jugnarain et al., 2022; Stagg et al., 2019).

Environmental restructuring can further support social opportunities and reduce automatic motivation barriers; an example would be to provide specialised clinics for under-served populations with holistic social support in addition to medical care. Providing training to healthcare workers in stigma reduction would continue to improve service environments for patients. Additionally, the provision of, and access to, harm reduction via sterile injecting equipment in appropriate environments remains a key gap identified from participants accounts of being unable to prevent reinfection (Askar et al., 2022). Persuasive messaging and incentives can be used to motivate people to engage in regular testing. Broader types of public health policy areas are depicted in Table 3; strengthening initiatives can support these interventions to become embedded into health services.

## Conclusion

In this study we interviewed people who are represented in quantitative research as at increased risk of reinfection (people who inject drugs, people who have been in prison (Hibbert et al., 2023)) and a participant who acquired HCV through sex. This research underscores the importance of accurate advice in prevention, testing, and treatment, and the social effect healthcare professionals and peers have in supporting patients and retaining people in care. Whilst participants reported improvements in care and knowledge from their first to subsequent reinfection(s), they report gaps concerning the continuity of care (e.g. from prison to release in community) and information provision (on all transmission risks and on future treatment eligibility) whilst receiving care. The barriers experienced were unique to each person and dependent on the setting - therefore this finding speaks to the need for holistic approaches, including harm reduction, mental health support, and addressing social stigma that impacts people’s mental health and community belonging. We found that the participants who had positive experiences of engaging in care, and of receiving peer support, felt enabled to either prevent further reinfection or to maintain regular HCV testing.

## Data Availability

All data produced in the present study are available upon reasonable request to the authors

## Author contributions

AS contributed to conceptualisation, data curation, formal analysis, investigation, methodology, project administration, writing-original draft and writing – review and editing. CSP contributed to data curation, formal analysis, writing - original draft, and writing - review and editing. MH contributed to conceptualisation, data curation, investigation, methodology, project administration, writing-original draft and writing – review and editing. RS contributed to conceptualisation, funding acquisition, resources, supervision, writing – review and editing. MD contributed to conceptualisation, funding acquisition, resources, supervision, writing – review and editing. LR, MR, JM contributed to resources, writing – review and editing, and LR also contributed to conceptualisation.

## Acknowledgements

We wish to acknowledge the support of the Hepatitis C Trust peers and employees who inputted on the study design and topic guide, including Stuart Smith.

We also wish to acknowledge support of NHS collaborators, including Duncan Cresswell and Sumita Verma at University Hospitals Sussex Foundation Trust, Mark Nelson at Chelsea and Westminster Hospital NHS Foundation Trust, Ashley Brown at Imperial College Healthcare NHS Trust, Stuart McPherson at Newcastle Upon Tyne Hospitals NHS Foundation Trust, and the supporting research staff.

We acknowledge the support of the NIHR Health Protection Research Unit on Blood Borne and Sexually Transmitted Infections (BBSTIs) steering committee: Professor Caroline Sabin (HPRU Director), Dr John Saunders (UKHSA Lead), Professor Catherine Mercer, Professor Gwenda Hughes, Dr Hamish Mohammed, Professor Greta Rait, Dr Ruth Simmons, Professor William Rosenberg, Dr Tamyo Mbisa, Professor Rosalind Raine, Dr Sema Mandal, Dr Rosamund Yu, Dr Samreen Ijaz, Dr Fabiana Lorencatto, Dr Rachel Hunter, Dr Kirsty Foster and Dr Mamooma Tahir.

## Ethical statement

NHS Research Ethics Committee and Health Research Authority approval was gained for the recruitment of NHS patients (IRAS: 331334). UK Health Security Agency research ethics and governance approval was obtained for recruitment via the Hepatitis C Trust (ID: 553). All participants gave informed consent.

## Data availability

The interview transcripts contain potentially confidential information and have therefore not been made publicly available but can be provided upon reasonable request to the corresponding author.

